# Glaucoma Detection and Staging from Visual Field Images using Machine Learning Techniques

**DOI:** 10.1101/2022.10.03.22280629

**Authors:** Nahida Akter, Jack Gordon, Sherry Li, Mikki Poon, Stuart Perry, John Fletcher, Thomas Chan, Andrew White, Maitreyee Roy

**Affiliations:** School of Optometry and Vision Science, UNSW Sydney, NSW 2052, Australia; School of Electrical and Data Engineering, University of Technology Sydney, NSW 2007, Australia; School of Electrical Engineering and Telecommunications, UNSW Sydney, NSW 2052, Australia; Faculty of Medicine & Health, University of Sydney, Sydney, NSW 2006, Australia; Department of Ophthalmology, Westmead Hospital, Sydney, NSW 2145, Australia; PersonalEYES, Sydney, NSW 2000, Australia

**Keywords:** Glaucoma, Machine learning, Deep learning, Visual field

## Abstract

**Purpose:** In this study, we investigated the performance of deep learning (DL) models to differentiate between normal and glaucomatous visual fields (VFs) and classify glaucoma from early to the advanced stage to observe if the DL model can stage glaucoma as Mills criteria using only the pattern deviation (PD) plots. The DL model results were compared with a machine learning (ML) classifier trained on conventional VF parameters.

**Methods:** A total of 265 PD plots and 265 numerical datasets of Humphrey 24-2 VF images were collected from 119 normal and 146 glaucomatous eyes to train the DL models to classify the images into four groups: normal, early glaucoma, moderate glaucoma, and advanced glaucoma. The two popular pre-trained DL models: ResNet18 and VGG16, were used to train the PD images using five-fold cross-validation (CV) and observed the performance using balanced, pre-augmented data (n= 476 images), imbalanced original data (n=265) and feature extraction. The trained images were further investigated using the Grad-CAM visualization technique. Moreover, four ML models were trained from the global indices: mean deviation (MD), pattern standard deviation (PSD) and visual field index (VFI), using five-fold CV to compare the classification performance with the DL model’s result.

**Results:** The DL model, ResNet18 trained from balanced, pre-augmented PD images, achieved high accuracy in classifying the groups with an overall F1-score: 96.8%, precision: 97.0%, recall: 96.9%, and specificity: 99.0%. The highest F1 score was 87.8% for ResNet18 with the original dataset and 88.7% for VGG16 with feature extraction. The DL models successfully localized the affected VF loss in PD plots. Among the ML models, the random forest (RF) classifier performed best with an F1 score of 96%.

**Conclusion:** The DL model trained from PD plots was promising in differentiating normal and glaucomatous groups and performed similarly to conventional global indices. Hence, the evidence-based DL model trained from PD images demonstrated that the DL model could stage glaucoma using only PD plots like Mills criteria. This automated DL model will assist clinicians in precision glaucoma detection and progression management during extensive glaucoma screening.

## Introduction

Glaucoma is a multi-factorial neuro-degenerative eye disease considered the second leading cause of blindness worldwide [1]. Till now, there are no definitive explanations for what causes glaucoma; but it is possible to detect and track the progression by continuing to monitor the VF loss and visible damage to the optic nerve head (ONH) of the retina.

Standard automated perimetry (SAP) has been the primary method for glaucoma diagnosis and progression follow-up [2]. The Humphreys Visual Field Analyser (HVFA) 24-2 and 30-2 SITA-Standard testing protocols are common perimeters used by clinicians to measure the severity of VF loss and grading glaucoma progression based on the Mills et al. criteria [3]. While being the gold standard for identifying vision loss, SAP can still be a complicated, time-consuming, and subjective process, as it is highly dependent on the clinicians’ experience and expertise [4].

The SAP test is a diagnostic metric used by clinicians to measure the sensitivity of functional vision in different parts of an individual’s VF. For glaucoma, the VF tests are essential as the clinicians can observe any peripheral vision loss before the patient experiences it. By assessing the reduced sensitivity or blind spots, they can grade the severity of the VF loss, and determine the appropriate management. Glaucomatous VF losses are dependent on the topology and anatomical distribution of the retinal nerve ganglion fibre bundles, which usually follow an arcuate pattern before entering the optic nerve. These fibres respect the horizontal raphe in the temporal retina. As a result, glaucoma gives rise to generalized and localized VF defects commonly formed in arcuate, altitudinal, nasal step, central, and paracentral scotomas, which respect the horizontal midline. For early glaucoma, the damage may present as partial arcuate, paracentral, and nasal step defects [5]. The VF defects can be widened by conforming to the ONH structural damages [6]. Therefore, an earlier diagnosis is crucial to halt the severe peripheral vision loss.

Nowadays, clinicians follow the most widely used glaucoma staging system suggested by Mills et al., which can classify glaucoma stages from 0 (no or minimal VF defect/ocular hypertension) to stage 5 (end-stage). The criteria were set based on the MD and PSD score, PD plot clusters, glaucoma hemifield test (GHT) or dB plots. Additionally, the staging system can grade glaucoma severity only from PD plots into four categories: early, moderate, advanced, and severe glaucoma.

The Mills grading system was developed based on the Hodapp-Anderson-Parrish (HAP) classification system, which also used VF parameters [7]. However, the HAP system could not classify the full range of glaucoma stages and was unable to separate the early and end-stage disease from minimal defects and blind patients [8]. Mills criteria provide a standardized grading system to use in clinical practice, which considers multiple parameters to classify the full range of glaucoma compared to the other grading systems: Advanced Glaucoma Intervention Study (AGIS) [9], Collaborative Initial Glaucoma Treatment Study (CIGTS) [10], Glaucoma Staging System (GSS) [11] and Enhanced Glaucoma Staging System (GSS2) [12].

Researchers have developed several ML algorithms utilizing VF images to differentiate glaucomatous eyes from normal eyes [13-19]; however, few ML studies have been done on automated glaucoma staging [20-24] using VF images. The outcomes of ML studies are promising for glaucoma differentiation from normal eyes using VF images and sometimes better than clinicians at recognizing complex patterns [19].

In this study, we aimed to investigate the DL model performance to differentiate normal VFs from glaucomatous VFs and classify glaucoma from early to advanced stages to observe if the DL model can stage glaucoma as Mills criteria using only the PD plots. The outputs of the DL models were compared with ML models trained on conventional VF global indices for the same classification.

## METHODS

### Datasets

In our retrospective study, a total of 265 reliable VF images were collected, 138 images from the UNSW Optometry Clinic and 127 images from the Centre of Eye Health (CFEH) (n = 265, normal eyes=119, glaucomatous eyes=146). The 265 PD plots and 265 numerical data were extracted from the Humphrey 24-2 SITA-Standard VF test from the HVFA (II-i 745i, Carl Zeiss Meditec, Inc. Dublin, CA); patients underwent for VF test from April 2011 to July 2022. The glaucoma data consists of 72 early, 43 moderate, 21 advanced and 10 severe patients. Due to the limited number of severe patients, we combined the severe patients with the advanced group. Expert clinicians label the glaucoma staging according to Mills et al. criteria. The VFs labeled ‘Preperimetric’ and ‘No glaucoma with RNFL losses’ were excluded from the glaucoma and normal group. The inclusion criteria for the VF test reliability were VFs with fixation losses of ≤ 20%, and false positive and false negative rates ≤ 15%. To be consistent with the PD plot pattern, the 10-2 and 30-2 SITA standard VF scans were excluded from the final dataset. The ethics committee of UNSW Sydney approved the relevant ethics for the data collection. The study followed the tenets of the Declaration of Helsinki, and written informed consent was collected from the patients to use the de-identified clinical data for research purposes.

### Power and sample size estimation

The power and sample size for the two classes (normal and glaucoma) and four classes (normal, early, moderate, and advanced) have been calculated. For two classes, the power was estimated using an independent t-test, with the required parameters: effect size d=0.50, α error = 0.05, group allocation ratio N2/N1= 1.2. For four classes, power was computed using F-tests, a linear multiple regression with the effect size, d=0.15 and α error = 0.05.

### Image pre-processing

To train DL models, only PD plots were cropped from the original VF images (Fig.1). The images were resized into 224×224 pixels for the input layer of the DL models. In the low-contrast images, the sharpness and contrasts have been enhanced.

**Figure 1:**
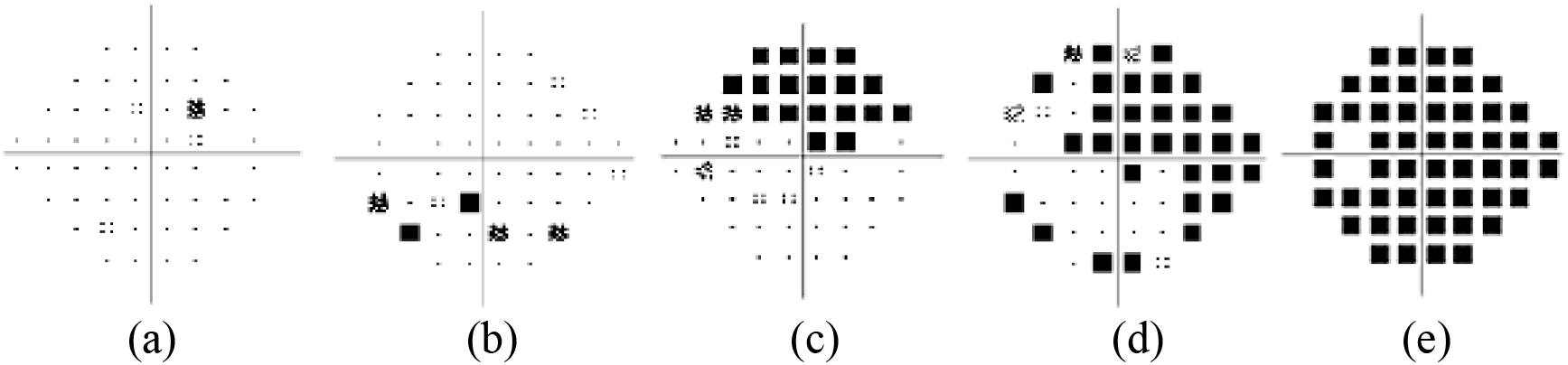
Pattern deviation plots of a) normal b) early glaucoma c) moderate glaucoma d) advanced glaucoma, and (e) severe glaucoma

### Deep learning models

Two pre-trained DL models, VGG-16 [25] and ResNet18 [26], were used to evaluate the classification performance of detecting and staging glaucoma using PD plots by classifying images into four classes: normal, early, moderate and advanced. ResNet and VGG are two popular CNN models that have shown remarkable progress in medical image analysis for smart medicine [27]. In the literature, these two models are commonly used in VF image prediction for glaucoma detection and progression grading [28-34]. VGG16 and ResNet18 consist of 41 and 71 layers and use 224×224-pixel size images in the input layers.

Having a small and imbalanced data in the classes, two combinations of images were used to train the DL models. In the first approach, the images were augmented and balanced in each group following the large image number group, i.e., normal=119. The image number was increased by performing a ‘horizontal flip’, ‘minus 5-degree rotation’, and ‘plus 5-degree rotation’ in the early, moderate, and advanced glaucoma group. Therefore, in the final image dataset, there were 476 images, 119 in each group. With a small dataset, we used a 5-fold CV in every DL approach to assess the generalizability of the DL models on unseen data and utilize the whole dataset for training.

In the first approach, the two pre-trained models were trained using a pre-augmented balanced dataset. In the CV, for each fold, one of the subsets was used as the test set, and the remaining subsets were placed together to make a training set. Hence, all the data got a chance to be trained in the model, and in the end, the evaluation scores were averaged across the 5 test subsets. Though image augmentation was done in the pre-processing step, during the model training, the MATLAB ‘imageDataAugmenter’ function was used for further data augmentation using three arguments: ‘RandXTranslation’, ‘RandYTranslation’ and ‘RandXShear’ to reduce the overfitting problem. The Stochastic Gradient Descent with Momentum (SGDM) optimizer (an iterative optimization algorithm to find the optimal result) with an initial learning rate of 0.0005 for ResNet18 and 0.00001 for VGG16 was used to train the models. The minimum batch size was set to 10, and a maximum epoch size of 20 was selected by monitoring the loss and accuracy of the training and validation dataset. Secondly, the two models were trained using the imbalanced image group (no pre-augmentation). Similarly, during model training, we used image augmentation using the arguments mentioned above to improve the model’s generalizability. This temporary datastore augments the images and doesn’t save them in the machine. The model updates the model parameters and discards the augmented images after the training. For a small number of images in the advanced group, the minibatch size was set to 5 and maximum epochs were set to 30. For ResNet18, the learning rate was 0.0005; for VGG16 the learning rate was 0.00001.

In the final DL method, the two pre-trained models, ResNet18 and VGG16, were used only for image feature extractors, and the extracted features were trained in a multiclass support vector machine (SVM) classifier to classify the images into four groups. The features of the final global pooling layer of the two pre-trained models were extracted to train the SVM classifier. The pooling layer pools the input features in the spatial locations, providing 512 features for VGG16 and 25088 for Resnet18. The output results were evaluated on the confusion matrix.

### Grad-CAM visualization

The DL-trained images were further investigated using the Gradient-weighted Class Activation Mapping (Grad-CAM) function [35] to visualize whether the models accurately classify the images by identifying the actual amount of VF loss of the PD plot. The Grad-CAM generates a heatmap for a specific class by highlighting the features of the image using the gradient of the prediction score from the SoftMax layer of the class with respect to the final convolutional layer of the model [35]. The heatmap transparently overlays on the predicted image, deep red refers to the peak value which has the greatest impact on the decision, and deep blue is the lowest class value.

### Machine learning models

The DL model results were compared with four popular ML classifiers trained on global indices extracted from VF images: MD, PSD and VFI. The RF classifier with a maximum depth of 5, multinominal logistic regression, k-nearest neighbors (k-NN, k=5) and support vector classifier (SVC) with linear kernel were used to train a total of 119 normal and 146 glaucomatous datasets. These ML classifiers were selected based on the previous literature outcomes on numerical multiclass data prediction [^30, 36, 37^]. The ML classifiers’ performance was observed on five-fold CV. Finally, the result of the best-performing ML model was compared with the DL model result based on the performance metrics derived from the confusion matrix (CM).

## RESULTS

The demographic characteristics of the data are shown in Table 1. Group differences were analyzed using an independent samples t-test and expressed as mean ± standard deviation (SD). All the extracted features in the two groups were statistically significant (p<0.05). A total of n=265 (normal 119 and glaucoma 146) data in two groups has greater than 95% power to detect a statistical difference using independent t-test for continuous variables. For four groups, n=265, the power was estimated 90%, and at least 27 samples are needed per group to identify a statistical difference using linear multiple regression.

**Table 1:** Demographic features of the study population

|  | Normal (n=119) | Glaucoma (n=146) | *P* value |
| --- | --- | --- | --- |
| **Age** (mean ± SD) | 41.74 ± 14.35 | 59.54 ± 16.91 | 0.000* |
| **MD** (mean ± SD) | -0.28 ± 1.30 | -7.03 ± 6.64 | 0.000* |
| **PSD** (mean ± SD) | 1.52 ± 0.42 | 5.86 ± 3.40 | 0.000* |
| **VFI** (mean ± SD) | 0.99 ± 0.01 | 0.82 ± 0.21 | 0.000* |

MD: mean deviation, PSD: pattern standard deviation, VFI: visual field index, \*significant

The pair representation of the features: MD, PSD and VFI of four groups are shown in Fig. 2 using distribution and scatter plots.

**Figure 2:**
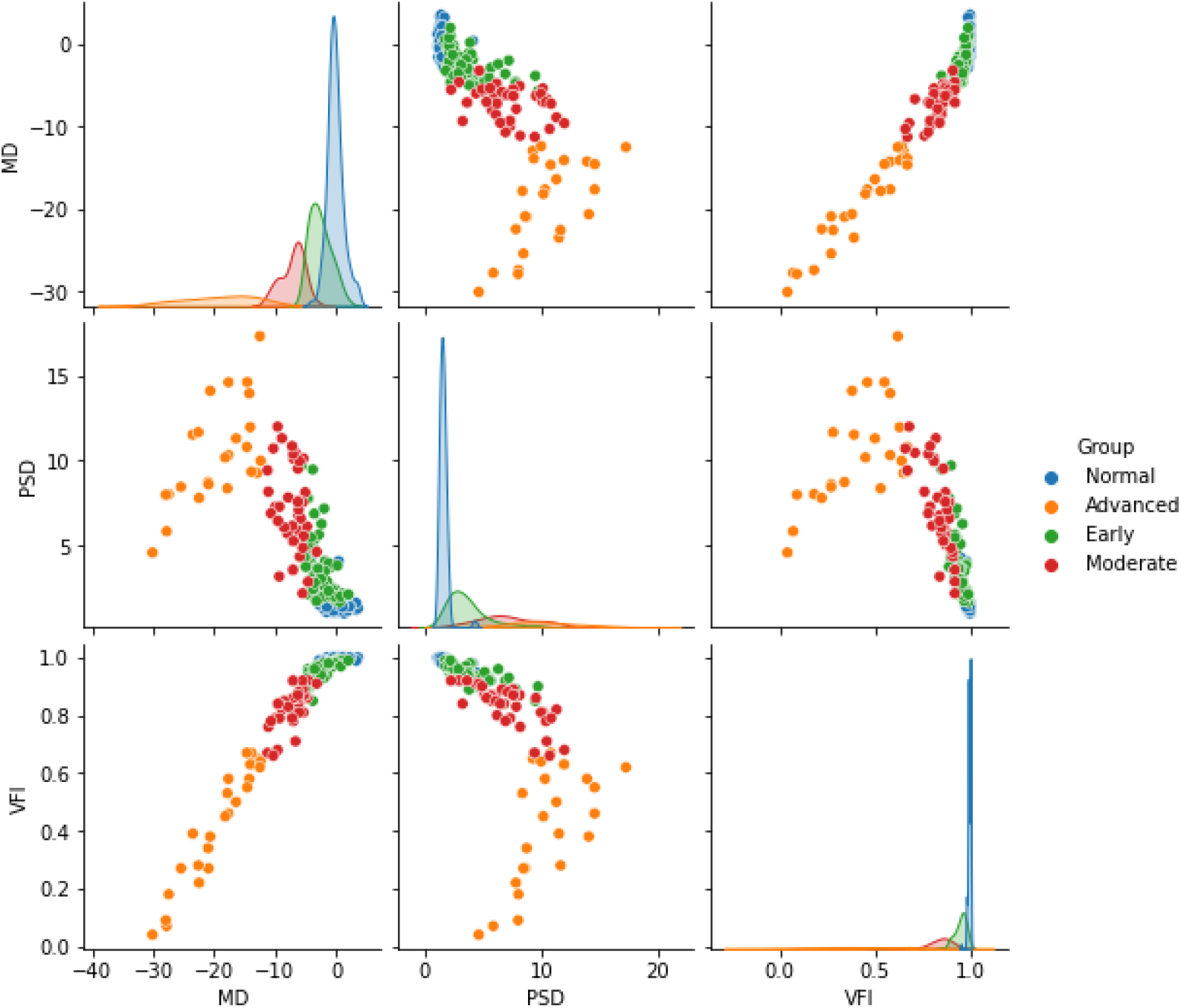
The pair scatter and distribution plot of the three global indices, MD, PSD and VFI, in relation to the normal and glaucoma group (early, moderate, and advanced).

The pair scatter and distribution plot in Fig.2 shows the features were overlapped for the early and normal groups. Besides, it is noticeable that the combination of MD and PSD pair can differentiate the normal to early group better than the MD, VFI or PSD, VFI pair. Though these MD, VFI or PSD, VFI pairs we can correctly discriminate the early, moderate, and advanced glaucoma group.

### DL model results

The following metrics have been used to estimate the DL and ML model performance and are defined as follows:

Sensitivity or Precision = TP/(TP+FN),

Recall = TP/(TP+FP),

F1= (2*TP)/(2*TP+FP+FN), and

Specificity= TN/(TN+FP),

Where TP is a true positive, TN is a true negative, FP is a false positive, and FN is a false negative. F1 score was considered as overall accuracy for multiclass classification.

The confusion matrices of the three approached DL models are shown in following Fig. 3.

**Figure 3:**
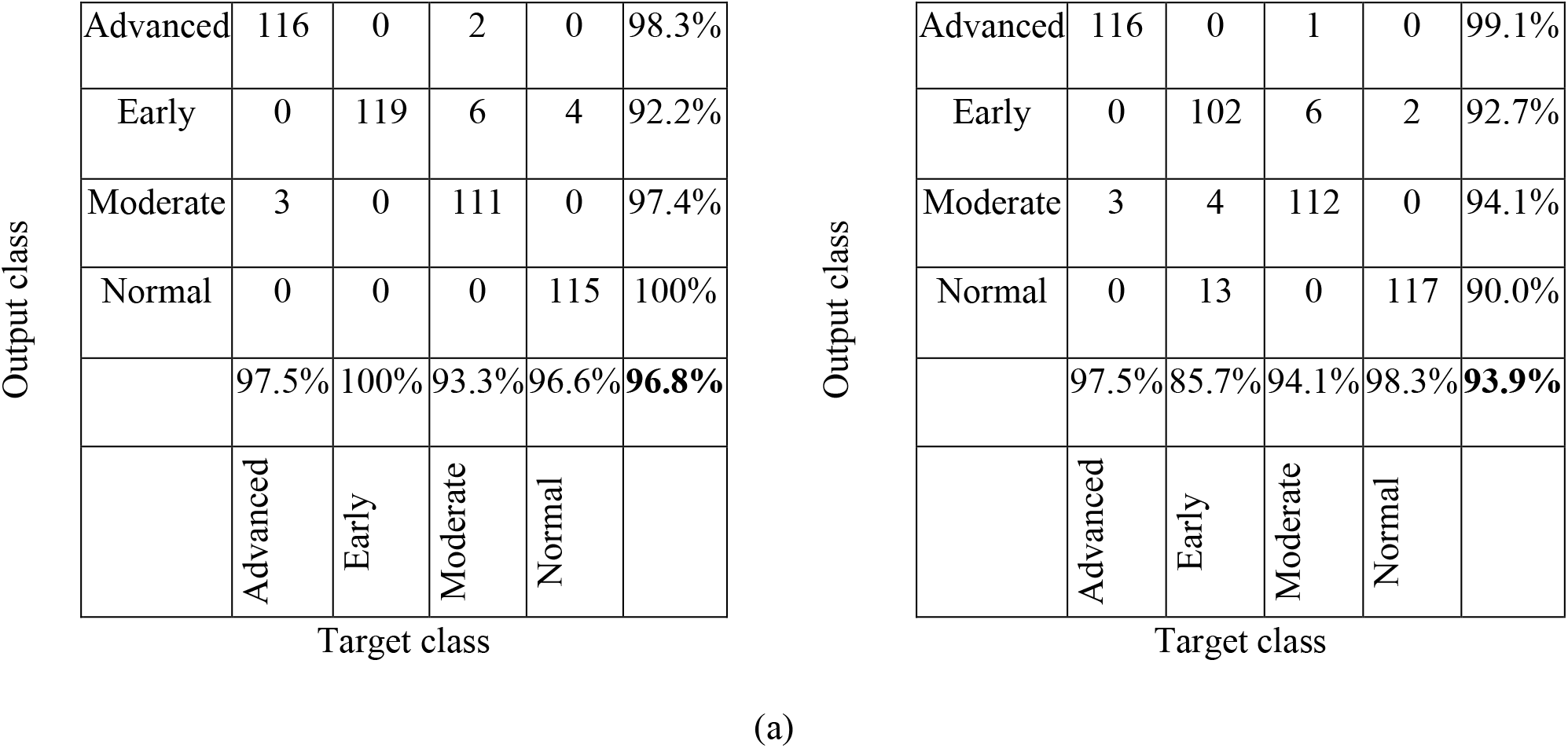
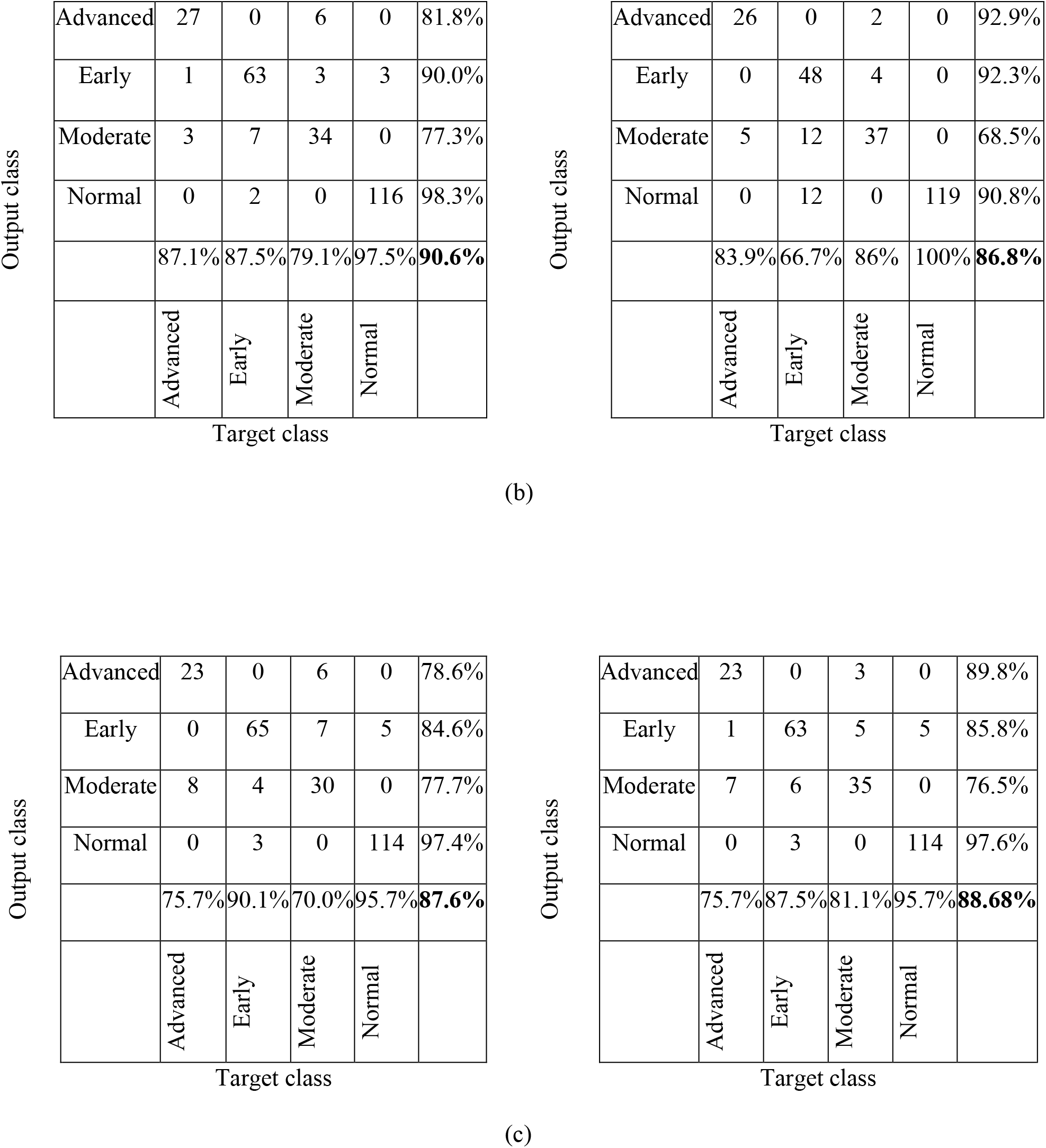
The confusion metrics of the ResNet18 (left) and VGG16 (right) model: classifying normal, early, moderate and advanced glaucoma using (a) pre-augmented balanced dataset (b) original imbalanced dataset (c) trained by a SVM classifier using extracted features using the original imbalanced dataset

In Fig.3 (a), the confusion matrixes showed both DL models performed well in classifying the four groups using the pre-augmented balanced dataset. The ResNet18 model achieved higher accuracy than the VGG16 model, with an overall F1 score of 96.8%. The other performance metrics (precision, recall and specificity) are shown in Table 3. The early cases were detected at 100%, and only four normal images were misclassified as early glaucoma. The normal eye images were further investigated, and it was that found the GHT was borderline for three images, and one eye was myopic with high-risk factors for glaucoma. In VGG16, though the false positive number for the early group was high (13) and overlapped with the moderate group, only two normal eye images were detected as glaucomatous eyes. The model showed similar performance for advanced and moderate glaucomatous images like ResNet18.

The performance of the DL models degraded when the original dataset (imbalanced) was used in two other methods (Fig.3 (b) and (c)). The detection rate of the glaucoma groups was significantly reduced, and the accuracy rate was highly varied between groups due to the imbalanced image number in the class. The VGG16 performed poorly on the dataset (Fig. 3(b) right). However, the two SVM classifier models performed almost similar when trained on extracted features from the two pre-trained models (Fig.3 (c)). Notably, the DL models distinguished the normal group from the glaucomatous group in three methods with high accuracy. For the original dataset in Fig. 3(b), the normal detection rate was 97.5% for ResNet18 and 100% for VGG16, proving that the PD plot trained from the DL model is a promising automated tool for separating glaucoma from normal eyes.

### Grad-CAM Visualization

The trained DL models were further investigated with Grad-CAM visualization, and a few resultant images are shown in Fig. 4.

**Figure 4:**
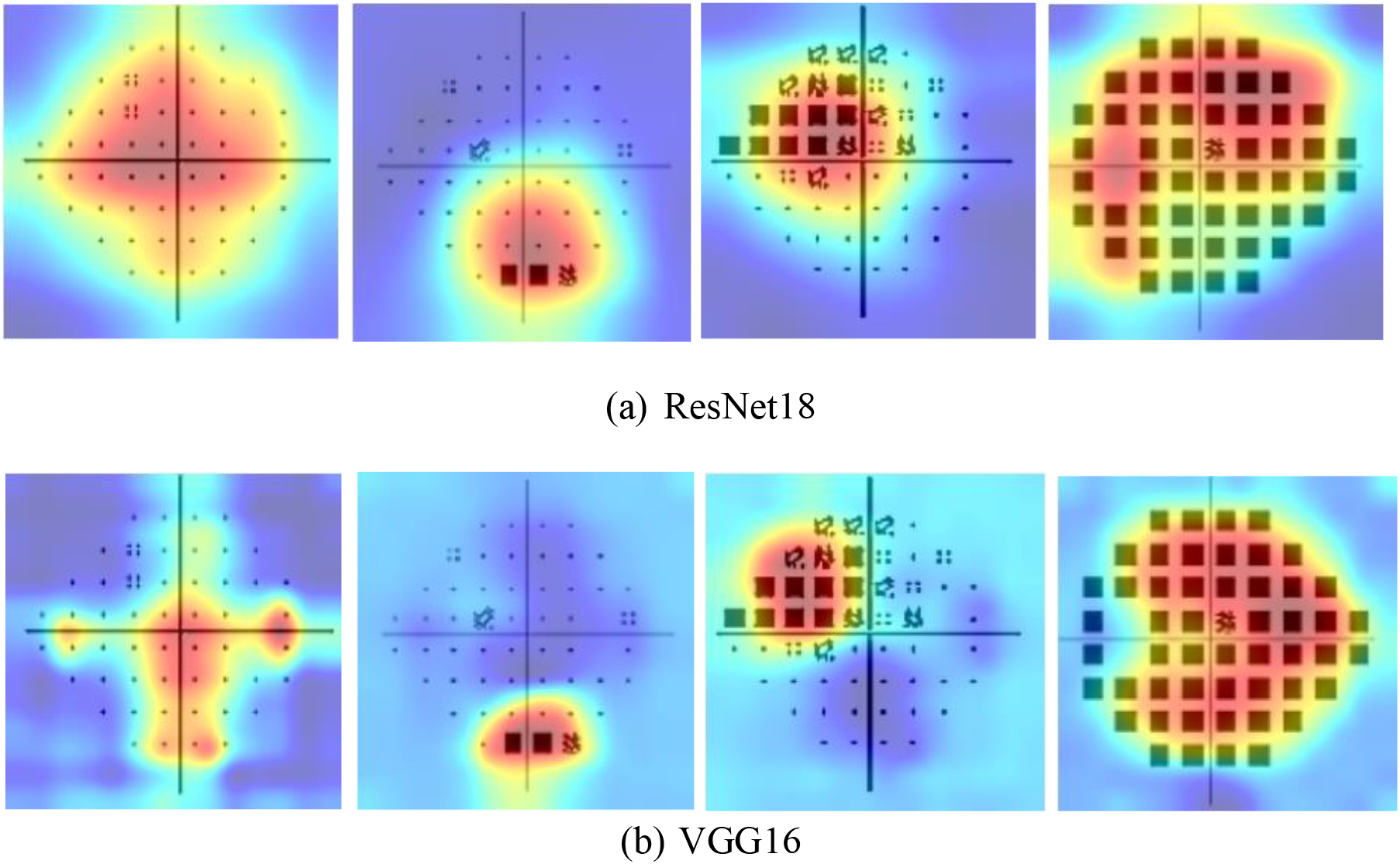
Generated heatmap for the visual explanation produced by Grad-CAM on pattern deviation plot scans (normal, early, moderate, advanced) from left to right in multi-class classification by (a) ResNet18 (b) VGG16. The Grad-CAM heatmap demonstrated that the ResNet18 and VGG16 models distinguished normal and glaucoma groups by localizing the affected regions in the visual field.

Fig.4 shows the heat maps of randomly selected categorical images of normal and glaucomatous eyes from each stage. Both DL models successfully identified the affected VF loss in the PD plots and distinguished normal VF images from glaucomatous images.

### Machine learning results

The RF model performed best among the ML classifiers. The F1 score for multinominal logistic regression was 92%, SVC (linear) model was 92% and K-NN was 90%. We have also trained the models with VFI and without VFI, and no effect was found in the classification result. Table 2 shows the performance summary of the RF model below.

**Table 2:** The performance metrics of the random forest model in classifying normal, early, moderate and advanced glaucoma using five-fold cross-validation.

| Group | Precision | Recall | F1-score | Number of data |
| --- | --- | --- | --- | --- |
| Normal | 97% | 97% | 97% | 119 |
| Early | 93% | 93% | 93% | 72 |
| Moderate | 93% | 95% | 94% | 43 |
| Advanced | 100% | 97% | 98% | 31 |
| **Overall** | **95.8%** | **95.5%** | **96.0%** | **265** |

Table 2 and 3 show that the performance of the best DL model (ResNet18) trained from a balanced dataset of PD plots and the best ML model (RF) trained from global indices is similarly performed for classifying the four groups: normal, early, moderate, and severe. The ResNet18 performed marginally better than RF mode, with precision: 96.98%, recall: 96.85% and F1 score: 96.8%. The DL model trained from the balanced dataset and ML model results proved that the PD plot could detect and stage glaucoma as the global indices. Moreover, it is also proven that in both cases, the ML model trained from VF data can separate glaucoma from healthy eyes and stage glaucoma from early to advanced stages based on Mills et al. criteria. However, compared with the original dataset, the DL model could not achieve as high accuracy as the ML model. Therefore, a large dataset with quantitative and qualitative analysis, including comparative assessment by the clinicians, is required to establish the result, which is our future study plan.

**Table 3:** The performance comparison summary of the DL and ML models for classifying normal, early, moderate and advanced glaucoma using five-fold cross-validation.

| Model | Precision | Recall | F1-score | Specificity | Number of data |
| --- | --- | --- | --- | --- | --- |
| ResNet18 (balanced data) | 96.98% | 96.85% | **96.86%** | 98.95% | 476 |
| VGG16 (balanced data) | 94.0% | 93.91% | 93.87% | 97.97% | 476 |
| ResNet18 (original data) | 87.76% | 87.79% | **87.77%** | 95.93% | 265 |
| VGG16 (original data) | 85.15% | 84.15% | 83.92% | 94.72% | 265 |
| ResNet18 (feature extraction) | 87.55% | 87.55% | 87.55% | 95.47% | 265 |
| VGG16 (feature extraction) | 88.68% | 88.68% | **88.68%** | 95.93% | 265 |
| RF model (global indices) | 95.8% | 95.5% | **96.0%** | 98.8% | 265 |

## Discussion

Our current study investigated three different approaches of DL models to detect and stage glaucoma using PD plots and compared them with ML models trained with global indices of VF images. The results demonstrated that a single PD image could classify glaucoma like the global indices: MD, PSD and VFI. Moreover, the accuracy of the deep learning model is improved when trained with a balanced dataset in multiclass classification.

Several studies have used ML classification algorithms to detect and stage glaucoma using VF images [15, 20, 24, 38]; however, very few used the PD plots for glaucoma detection [19, 34, 39-41], and we did not found any studies on glaucoma staging. Recently, Shin et al. used wide-field OCT-based DL classification to categorize the images into five groups: normal, pre-perimetric glaucoma, early, moderate, and severe glaucoma. The model achieved high accuracy for separating the normal group from the glaucoma group but showed difficulties in staging glaucoma with 72.69% accuracy [28]. Another study by Huang et al. used a 30-2 total deviation plot of VF images to train a fine-graded DL model to classify the images into five categories: clear VF, mild, moderate, severe, and diffuse VF defect. The model achieved an AUC of 0.93 with an accuracy of 85%. Zhen et al. trained DL models with fundus images to detect the glaucoma severity and achieved an accuracy of 85.29%. Though increasing the accuracy of the glaucoma staging still needs DL model improvement with diverse datasets, these studies’ strength is that the DL models were trained on a large training dataset. In contrast, in our study, the DL model with the original dataset achieved an F1 score of 87.8%, and with features extraction, the F1 score was 88.7% to classify the images into normal, early, moderate, and advanced groups. For detecting glaucomatous VF images, it achieved the AUC: 0.99 with an accuracy of 98.1%, and for staging (early, moderate, and severe), the DL model achieved an F1 score of 85.8%, which is promising for using PD plots for automated glaucoma diagnosis and progression grading.

In this study, we used the Mills et al. grading system as the foundation for our glaucoma grading. Although there is still no clinically accepted gold standard, this is the most popular grading system in clinical practice [42]. This criterion considers the MD, PSD, GHT, and PD plots to classify the full range of glaucoma from early to end-stage disease. We focused on using the PD plots for training data, as this plot best highlights any localised defects by taking the difference of the observed threshold at one point from the average threshold hold across the entire VF. Thus, this eliminates any field defects caused by cataracts, giving a more accurate representation for glaucomatous loss [43].

In clinical practice, diagnosing glaucoma involves a combined analysis and multimodal assessment of the structural and functional glaucomatous damage via OCT, VFs, and stereoscopic fundus images [44]. While characteristic glaucomatous VF defects may appear in some glaucoma patients, atypical patterns can still occur due to the various VF patterns that can be presented within glaucoma. As a result, this becomes a complex process to interpret, diagnose and stage glaucoma. Glaucoma diagnosis and staging not only involve interpretation of the PD maps, but also of other features such as global indices, which typically form part of the criteria in established glaucoma staging systems such as that by Mills et al. However, our study revealed that the DL model could use PD maps alone to detect glaucoma and stage glaucoma severity with the same performance of these trend analysis values. Therefore, the PD map can be a novel clinical imaging feature to train artificial neural networks for automated glaucoma detection and classification. Glaucoma diagnosis is a complex, time-consuming process requiring trained and skilled practitioners to interpret results. Thus, by implementing this into clinical practice, glaucoma detection and classification from just PD images may potentially improve the efficacy of clinicians as well as reduce the patient burdens associated with invasive clinical procedures. Moreover, an artificial neural network can be useful in the early detection of glaucoma, prompting early interventions to prevent progressive vision loss and the negative impacts on the patient’s quality of life.

Our study also demonstrated the visual explanation of the DL model in decision-making on the images based on the correctly localized affected VF loss. This refers to practitioners relying on our DL systems that provide automated predictions and visual explanations. The DL model can discriminate patterns between normal and glaucoma and the glaucoma severity in the VF images. Ultimately, this explainable DL-based glaucoma detection and staging will help the eye care professionals and clinicians to trust the DL algorithm and rely on the clinical decisions. Our study has some limitations. Firstly, the limited amount of training datasets restricts the validity of our DL model. Large amounts of data are required to train these ML and DL algorithms to be “well-phenotyped” to the disease and optimize their diagnostic performances. As our clinical data were obtained from the CFEH, a secondary referral centre, and the UNSW Optometry clinic, most of our dataset consists of normal and early eyes (who may also be glaucoma suspects) with limited amounts of moderate and advanced cases. Our ML models showed difficulties in differentiating the early and moderate glaucoma stages. In clinical practice, this is a vital delineation that dictates the management plan, whereby early glaucoma can be initially managed by an optometrist.

In contrast, advanced glaucoma is not suitable for collaborative care and requires referral to an ophthalmologist [45]. Although we have collected data from two eye care centres, future directions involve incorporating data from tertiary referral centres such as eye hospitals and ophthalmologic clinics, which may present more availability of moderate to severe glaucoma cases and improve the performance of the ML model. Secondly, including more data from these centres may reduce ethnicity bias. In our current clinical dataset, there was an increased number of VFs obtained from Caucasian and Asian patients. As a result, our study cannot truly reflect the disease into the broader population or explore the epidemiological effects of ethnicity on glaucoma. It is important to note that obtaining large amounts of high-quality data may also be associated with time constraints and technological difficulties and ultimately be considered financially unfeasible [46].

Thirdly, with glaucoma being a multifactorial disease, we could not correlate and validate other diagnostic features and risk factors of glaucoma such as RNFL, cup-disc ratios, IOPs, age and family history. Furthermore, our study primarily focused on distinguishing between normal and glaucomatous defects from a database that was previously grouped as glaucoma or healthy. Our study is limited to differentiating glaucoma from other ocular pathology that may present with the same VF loss patterns.

Fourthly, our study is entirely dependent on PD plots only extracted from HVFA 24-2 SITA Standard VFs. The 30-2 protocol has also been implemented to investigate glaucomatous VF loss, employing a 76-point test grid over the central 30 degrees of the VF. Hence, there is a potential of missing any mid-peripheral glaucomatous scotomas. Though, the 24-2 testing strategy has been shown to provide comparable information to that provided by the 30-2 protocol in more than 95% of cases, with advantages of shorter test duration and less variability [47]. Thus, VF printouts under the 30-2 testing strategy could be considered as part of the dataset, which can ultimately increase the training dataset and strengthen the performance of the artificial neural network.

In conclusion, our proposed DL-based glaucoma detection and staging using PD plots showed promising and comparable performance in contrast to conventional functional parameters. Glaucoma diagnosis involves a complicated, time-consuming, and subjective process dependent on the clinician’s experience and expertise. Hence, with the high diagnostic accuracy of the DL model in detecting glaucoma, we can consider its application within a clinical practice to assist in yielding more accurate glaucoma diagnosis and, in particular, early detection of glaucomatous VF loss. Although the DL results of this study are promising for glaucoma detection and classification, further data collection and comparative study are warranted to train the algorithm with a larger number of VF datasets to validate the results and make it clinically useful and reliable.

## Data Availability

All data produced in the present study are available upon reasonable request to the authors

## Acknowledgement

We would like to thank Dr Kathleen Watt, Clinic Director of UNSW Optometry clinic, for helping us with the data collection. We also want to thank the clinicians of CFEH, UNSW Sydney, for assisting in the data collection. Guide Dogs NSW/ACT funds clinical services at the CFEH. Guide Dogs NSW/ACT had no role in study design, data collection and analysis, publication decisions, or manuscript preparation.

